# The factors associated with stillbirth in urban Vadodara: A population based cross-sectional study, Vadodara Municipal Corporation, Gujarat, India

**DOI:** 10.64898/2026.09.30.26364463

**Authors:** Brijesh Rameshbhai Gadhethariya, Bharati Makwana, Jivraj Damor, Priyash Sheth, Urvish Patel

## Abstract

**Background:** Stillbirths are a neglected problem in India, and the data we have are often unreliable. This study aimed to assess stillbirth record quality in an urban setting and understand the experiences, health impacts, and perceived causes from both families and health workers.

**Methods:** A mixed-methods study was conducted in Vadodara Municipal Corporation during January– December 2023. All reported stillbirths (≥28 weeks or ≥1000 g) were reviewed. Families were interviewed at home using a standardized questionnaire, with in-depth interviews conducted with 9 families and 17 health workers (doctors, ANMs, ASHAs). Quantitative data were analysed using Excel and Jamovi; qualitative data were analysed thematically.

**Results:** Of 61 stillbirths reported by VMC, quality audits revealed 7 errors (duplicates/misclassifications) and 2 data entry errors, yielding a true stillbirth rate of 1.7 per 1000 total births. The eastern zone recorded the most cases (21), while the southern zone had the highest rate (4.1 per 1000). Most deliveries occurred in government hospitals (64%). Common reported causes were intrauterine foetal death on routine ultrasound (26%) and premature delivery (19%). Families reported delayed care access, undertrained staff, poor facilities, and challenges accessing government scheme benefits. Health workers identified preventable factors (hypertension, diabetes, anaemia, meconium aspiration, growth restriction, birth defects) and non-preventable factors (extreme prematurity, multiple pregnancy).

**Interpretation & conclusions:** Stillbirth data in Vadodara have serious quality problems which can be fixed. Many stillbirths could be avoided with better pregnancy monitoring, better communication, stronger referral systems and more awareness in the community.

## INTRODUCTION

Stillbirths, an important indicator of the quality of care during pregnancy and childbirth, occur alarmingly frequently in many countries. Although progress has been made in reducing the global stillbirth rate, gains are uneven, with substantial inequities persisting among regions and countries.(United Nations Inter-agency Group for Child Mortality Estimation (UN IGME) 2023)

Stillbirth is defined as the complete expulsion or extraction of a foetus that died prior to this expulsion/extraction, at ≥22 completed weeks of gestation.(World Health Organization 2019)

This is a major global health problem affecting 7,000 families daily and carries emotional, social, and economic consequences.(Lawn et al. 2016) Currently, 98% occur in low- and middle-income countries (LMICs), with India having the highest number (~592,100 deaths/year) and a WHO-estimated rate of 22 per 1,000 total births.(Lawn et al. 2016; Altijani et al. 2018)

Many social and biological factors are associated with stillbirth; appreciating these factors will greatly impact reducing its rate. Proven risk factors include socioeconomic deprivation (female illiteracy, low socioeconomic status, rural residence, caste, women not in paid employment, and religion) and addictions (e.g., tobacco smoking), as well as adverse pregnancy outcomes and birth characteristics.(Altijani et al. 2018) Antepartum causes were more common (68%) than intrapartum (32%), with foetal (48%) and maternal (44.7%) causes unidentifiable in about half of cases.(Newtonraj et al. 2017) Maternal conditions such as severe anaemia, hypertension, diabetes, HIV, eclampsia/convulsions, and foetal distress critically affect pregnancy care, while delivery mode (normal vaginal or assisted/instrumental) and the birth attendant influence intrapartum care.(Singh et al. 2019)

Globally, over 5,000 babies are stillborn daily at ≥28 weeks’ gestation, totalling 1.9 million annually.(United Nations Inter-agency Group for Child Mortality Estimation (UN IGME) 2023) Many births are inaccurately assessed, and key data are improperly recorded because data collection tools may be unavailable or non-standardized.

Health information is a fundamental management tool with a key impact on societal progress. Its objective is to provide reliable, relevant, up-to-date, adequate, timely, and reasonably complete information for health management at all levels. However, obtaining such information remains difficult; recorded data from some countries suffer from quality issues due to non-standardized definitions, underreporting, or misclassification of stillbirths. Strengthening data collection and improving data quality within health management information systems (HMIS) and household surveys are key to addressing these challenges.

Despite their large number, efforts to measure stillbirths lag behind in many countries; without systematic monitoring of rates, numbers, and causes, these tragedies will persist. Research was needed on stillbirth reporting and data quality to achieve broader maternal and newborn health goals.

### Problem or Issue

Stillbirth remains under-recorded and poorly audited in urban India, obscuring the true burden, contributing causes, and gaps in the care pathways that families experience around the event.

### What is Already Known

India has the world’s highest stillbirth burden, linked to socioeconomic deprivation, maternal illness, and delayed care. However, routine surveillance data quality and local care-pathway experiences are rarely examined together.

### What this Paper Adds

This population-based mixed-methods audit of an entire urban corporation corrects reporting errors to reveal the true stillbirth rate, maps family care pathways, and identifies preventable causes from provider and community perspectives to guide targeted action.

## AIM AND OBJECTIVES

1. To review the reported stillbirth data regarding classification and stillbirth rate among stillbirths in Vadodara Municipal Corporation area.
2. To assess care pathways and health problems experienced by family during stillbirth.
3. To identify associated factors and causes for stillbirth from community and health care provider perspective

### Participants, Ethics and Methods

This mixed□methods study employed an exploratory cross□sectional design and was conducted within the Vadodara Municipal Corporation (VMC) area from January to December 2023, with an overall study duration of 18 months. The study population comprised all families who experienced a reported stillbirth (defined as foetal death at 28 or more completed weeks of gestation or with a birthweight of 1000 grams or more when gestational age was unavailable) as per the VMC health management information system, along with the healthcare providers involved in their care. Families residing within the VMC jurisdiction were included, while those who had migrated out of the area during the study period were excluded. To minimise recall bias, household visits were conducted within approximately 21 days of the stillbirth event, with the assistance of local community health workers, including ASHAs and FHWs. Data were collected using two parallel approaches: for the quantitative component, a semi□structured, pretested questionnaire in the local vernacular was administered to family members after obtaining written informed consent, with pretesting carried out at the Ektanagar UPHC; for the qualitative component, in□depth interviews lasting 45 to 50 minutes were conducted with a purposive sample of nine affected families and seventeen healthcare providers (including medical officers, ANMs, and ASHAs) using a predesigned interview guide. All in□depth interviews were audio□recorded with participants’ consent and subsequently transcribed, translated, and coded on the same day, with non□verbal communications and observations also noted. All offline data were digitised using Microsoft Office applications for subsequent analysis.

Quantitative data were entered and analysed using Microsoft Excel 2021, Microsoft Word 2021, Visual Anthropac 1.0 (for free□listing and pile□sorting exercises), and Jamovi 2.3.21 software, with results expressed as proportions and percentages. For qualitative data, demographic characteristics and reported causal factors were first categorised into predefined groups (antepartum causes such as pre□conception and antenatal care issues, and intrapartum causes during labour and birth), and these were presented as frequency tables, bar charts, and pie charts. Concurrently, the care pathway evaluation data were subjected to thematic analysis using manual content analysis, from which emerging themes, codes, and verbatim quotes were systematically extracted and reported. A strict quality assurance and quality control protocol was applied throughout data entry and analysis, using cross□verification files to ensure accuracy. Ethical approval was obtained from the Institutional Ethics Committee for Human Research – PG Research (IECHR□PGR), and all participants were fully informed about the study procedures before providing written informed consent; privacy and confidentiality were strictly maintained, with all data stored in password□protected Excel files. Standard operational definitions were applied throughout, including the WHO□based stillbirth definition, birthweight classification (normal: >2500g; low birthweight: 1500□2499g; very low: 1000□1499g; extremely low:

<1000g), neonatal death, stillbirth rate per 1000 total births, young primipara (first delivery at ≤15 years), elderly gravida (first pregnancy at ≥35 years), and IFA intake per need calculated as the ratio of actual IFA days consumed to the ideal requirement based on gestational age.

## RESULTS

### Objective 1: To review the reported stillbirth data regarding classification and stillbirth rate among stillbirths in Vadodara Municipal Corporation area

The Vadodara Municipal Corporation (VMC) reported 61 stillbirths after details collected from the HMIS reporting by all the UPHC under them. We visited and collected details from each family members experienced stillbirths reported by VMC. Based on collected data, we reviewed the reported data and done comparative analysis which is as described in Table 1.

**TABLE 1:** COMPARATIVE ANALYSIS AFTER REVIEWING REPORTED STILLBIRTHS IN VADODARA MUNICIPAL CORPORATION AREA.

| Topic | Total reported stillbirths (n =61) |  | Description of discrepancies |
| --- | --- | --- | --- |
| Reporting time | Jan - March 2023 (n = 12) | April - December 2023 (n = 49) |  |
| Line list provided | 12 | 48 (-1) | 1 entry missing / over counting in Adarshnagar UPHC |
| Duplicate entry | 11 (-1) | 47 (-1) | 2 duplicate entries by UPHC Kishanwadi (Kajal Vijay Machi) and UPHC Makarpura (Mittal Rohit) in both groups respectively |
| Combined data entry | 11 | 48 (+1) | 2 entries combined in 1 line by Ektanagar UPHC and counted only 1 stillbirth. |
| Misclassified reporting | 11 | 46 (-2) | 1 neonatal death was reported in Shiyabaug UPHC as stillbirth.<br>1 abortion was reported as stillbirth in UPHC Sawad |
| Twin death | 12 (+1) | 46 | 1 Twin death in Kishanwadi UPHC counted as 1 stillbirth. |
| Final total stillbirths | (n = 58) |  |  |

Addressing these misclassifications and reporting errors at the UPHC level will enhance the accuracy of stillbirth data before it is reported to higher authorities.

Principal informant regarding this unexpected occurrence was mother 70.68% (n = 41) either independent or with other family member followed by father 34.48% (n = 20). Other family members (Grandmother, Aunty) contribution is minimum from 1.7% (n = 1) to 5.1% (n = 3). The other categories have minimal representation, indicating that the primary caregivers or respondents were predominantly parents.

As per Table 2 regarding zonal distribution of stillbirth reporting, east zone reported highest stillbirth (n = 21) followed by south zone (n = 18), North zone (n = 14) and very few stillbirths reported from Western Zone (n = 5) of Vadodara Municipal Corporation, which was statistically significant by Chi-square Goodness of Fit (χ2 = 10, P = 0.019). although stillbirth rate of zone shows different scenario having Highest stillbirth rate 4.1 per 1000 total births in South Zone followed by East Zone (2.3 per 1000 total births), North Zone (1.2 per 1000 total births) and West Zone (0.5 per 1000 total births). This shows good pregnancy outcome in east zone as compared to south zone, but both zones had stillbirth rate higher than average stillbirth rate (1.7 per 1000 total births) of Vadodara Municipal Corporation. While North zone and West zone had lower stillbirth rate showing good pregnancy outcome in that zone.

**TABLE 2:**
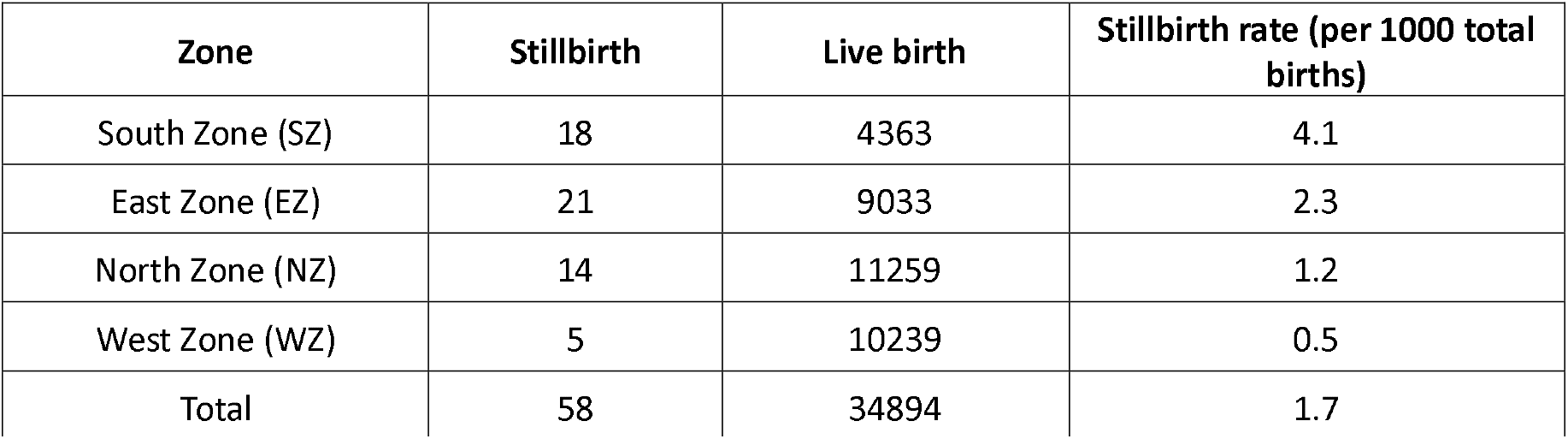
ZONE WISE DISTRIBUTION OF STILLBIRTH, LIVE BIRTH AND STILLBIRTH RATE (N = 58)

| Zone | Stillbirth | Live birth | Stillbirth rate (per 1000 total births) |
| --- | --- | --- | --- |
| South Zone (SZ) | 18 | 4363 | 4.1 |
| East Zone (EZ) | 21 | 9033 | 2.3 |
| North Zone (NZ) | 14 | 11259 | 1.2 |
| West Zone (WZ) | 5 | 10239 | 0.5 |
| Total | 58 | 34894 | 1.7 |

Out of 40 UPHC, 22 UPHC reported Stillbirth. SAWAD UPHC reported highest stillbirth (n = 7) followed by PANIGATE UPHC (n = 5), EKTANAGAR UPHC (n = 4) and VADSAR UPHC (n = 4). Remaining all UPHC reported <=3 stillbirths. We didn’t find specific reason for high stillbirth in this area.

The highest number of stillbirths occurred in June, with a peak of 9 cases. In June, the stillbirth rate reached its highest point at 3.15 per 1000 live birth. This indicates that a significant proportion of pregnancies during that month resulted in stillbirths. Conversely, August had the lowest stillbirth rate, recorded at 0.30 per 1000 live birth.

The majority of mothers were aged 21–30 years (74.1%), 6 mothers (10.34%) were found to be in high-risk group (<20 years is YOUNG PRIMI and > 35 years is ELDERLY GRAVIDA) according to RMNCAH+ A criteria of high-risk Pregnant women. The majority of the women (48.3%) are in their first pregnancy, with a progressively smaller percentage of women having had more pregnancies. Almost half 31 (53.4%) women were housewives and most women were from Hindu 44 (75.9%) and General caste 29 (50.0%). Majority of families were staying in Pucca houses 49 (84.5%).

Approximately one-third (20, 34.5%) of stillborn babies have a normal birth weight, while another third (23, 39.7%) fall into the Low-Birth-Weight category based on classification criteria. Interestingly, only three (5.2%) stillborn babies exhibit a gestational age of less than 28 weeks combined with a birth weight exceeding 1000 grams. Additionally, six (10.3%) stillborn babies weigh less than 1000 grams (classified as Extremely Low Birth Weight) despite being more than 28 weeks into gestation. Notably, no significant gender differences were observed among the stillborn cases.

Compared with LMP-based calculation, 3 stillbirths (5.2%) were misclassified by gestational age in the verbal autopsy records. These 3 stillbirths were from UPHC Sawad, Gotri and Fatehganj.

We found out that among the place of delivery Government Hospitals 37 (63.8%) dominate the delivery followed by private hospitals 15 (25.9%) and trust hospitals 6 (10.3%). Among the government hospital, SSG Hospital 28 (48.3%) was contributing major part of delivery due to tertiary health care level of government hospital.

The Figure 1 provides an analysis of the causes of stillbirth based on verbal autopsies, categorized into those occurring before birth (Antenatal, N = 50 (86.2%)) and during birth (Intranatal, N = 8 (13.8%)). The majority of stillbirths (86.2%) were identified as happening during the antenatal period. IUFD (intrauterine foetal death) on routine investigation (15, 25.9%) and premature delivery (11, 19.0%) were leading cause for stillbirth reported followed by low birth weight (8, 13.8%), High risk ANC (4, 6.9%), Placental Abruption (4, 6.9%), Meconium Aspiration (4, 6.9%), High BP during Labour/ Preeclampsia (4, 6.9%) and complications from twin pregnancies (3.4%). A few cases had no identifiable cause (6.9%).

**FIGURE 1:**
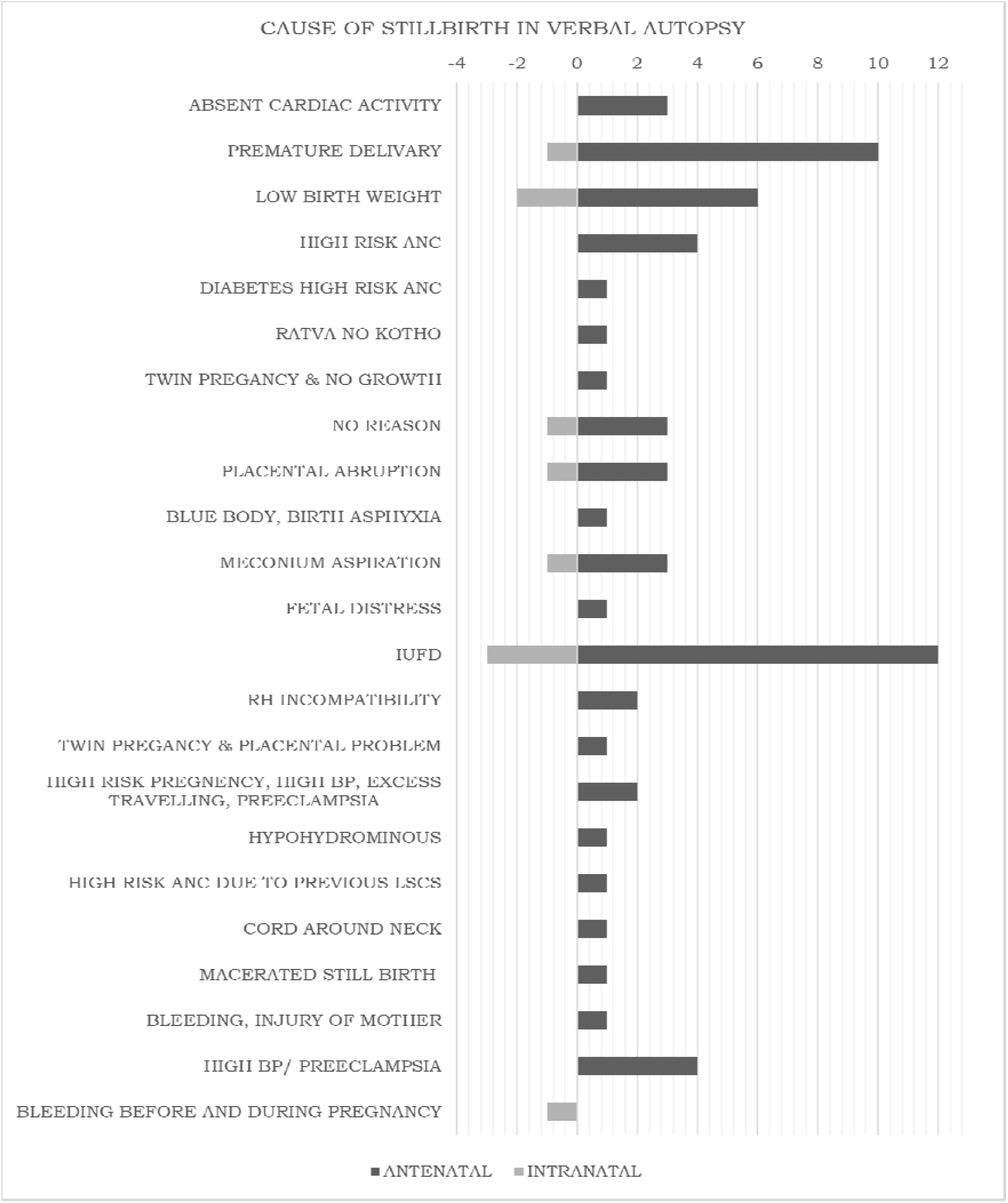
CAUSES OF STILLBIRTH IN VERBAL AUTOPSY (N = 58)

Out of all reported stillbirth, we find out 4 reporting errors which should be addressed properly because they were affecting the parameter like male: female ratio and classification of stillbirth based on gestational age. Out of 4, one had gender error by UPHC Chhani and other 3 have errors in gestational age calculation reported in verbal autopsy by UPHC Sawad, Gotri and Fatehganj.

### Objective 2: To assess care pathways and health problems experienced by family during stillbirth

To assess the health problems experienced by family during stillbirth was reviewed as antenatal care report in three categories (A) Symptoms observed during Pregnancy, (B) delivery Outcomes and (C) Investigations and care provided during the pregnancy. These 3 were described as following.

#### Antenatal Care Report (N = 58)

##### Symptoms Observed

Based on symptomatic observation by family member, we found out that 3 (5.2%) mothers exhibited pale and white appearances, 3 (5.2%) mothers had swelling on face and feet and 4 (6.9%) mothers had swelling over feet only. Other symptoms like breathlessness while doing Household Chores, Persistent Headache, fits without fever and fits prior to pregnancy was reported vary less. Mild fever during pregnancy was reported in 4 (6.9%) cases. out of these 4, two mothers had cough and 1 mother had rash associated with fever. Out of 58 mothers, 8 (13.8%) mothers had started bleeding per vaginally and 11 (19.0%) mothers had ruptured bag of water before the onset of labor pain. 54 (93.1%) mothers had stopped foetal movement before onset of labor pain. Baby got stuck in 2 (3.4%) mothers during delivery. One case of gestational hypertension and one case of hypothyroidism was found. Twelve (20.7%) mothers were taken to other hospital just before delivery.

##### Delivery Outcomes

Out of the 58 deliveries studied, 35 were performed through Normal Vaginal Delivery (NVD), while 23 were Caesarean Sections (LSCS). Among the LSCS cases, three mothers experienced malpresentation: two with breech presentation and one with shoulder presentation.

##### Investigations and Care Provided

Investigation and care provided during pregnancy included 3 parameter which is blood testing, USG and Blood pressure at the time of delivery. Among the 58 mothers, seven (12.1%) were found to be anaemic, one (1.7%) had diabetes, and two (3.4%) carried the sickle cell trait with normal haemoglobin levels. Additionally, one (1.7%) mother was reported to be hyperthyroid with anaemia. Eleven (19.0%) mothers experienced high blood pressure during delivery, while only one (1.7%) mother had low blood pressure. Fifty-four (93.1%) mothers underwent anomaly scans by ultrasound during pregnancy, revealing normal foetal growth. While one (1.7%) mother found, Oligohydramnios, one (1.7%) had pyelonephritis and growth retardation, one (1.7%) had cord around neck and one (1.7%) had congenital kidney disease and polyhydramnios.

According to the data, pregnant women who didn’t seek care or attended only 1, 2, or 3 ANC (antenatal care) visits at government hospitals were more likely to receive care from either Private Hospitals or Trust Hospitals. The majority of women received more than 3 ANC visits across any of the described facilities. Additionally, a few pregnant women received care from more than one setting, combining services from either Government and Private Hospitals or Government and Trust Hospitals.

To assess the care pathway, we made a flow chart of referral and contacts made by family during the delivery.

Now to describe care pathway in details, We divided it in two parts (I) delivery conducted in 1^st^ contacted hospital and (II) delivery conducted after ≥ 2^nd^ contacted hospitals. (Figure 2)

**Figure 2:**
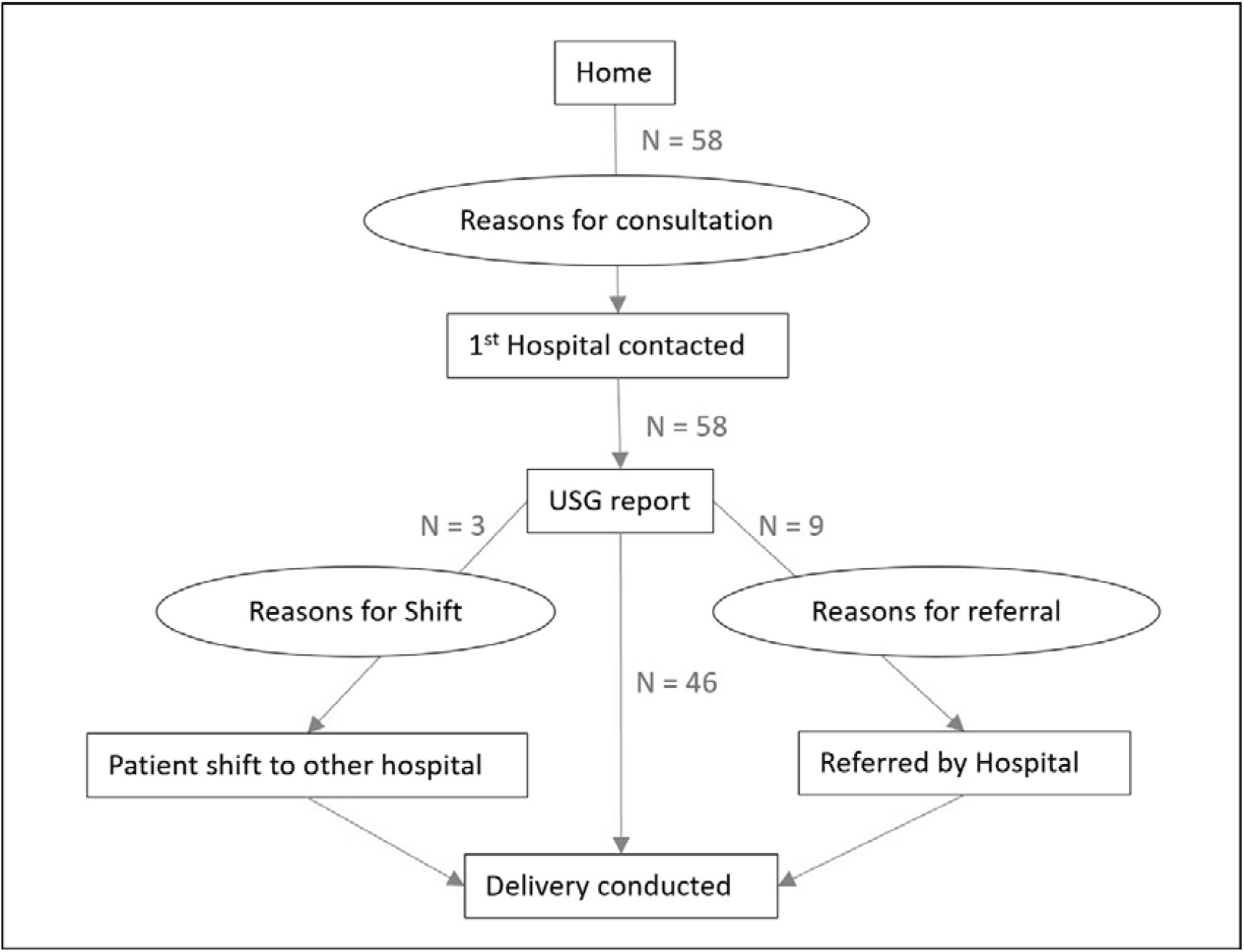
Flow chart of care pathway about referral and contacts made by family during the delivery (N = 58)

#### Delivery conducted in 1^st^ contacted hospital (N = 46)

Each and every care pathway begin with the consultation at hospital 1 ^st^ time and ends with outcome of health problem. Here care pathway starts with reason for consultation at hospital and ends to Stillborn by different delivery methods adopted. Reason for consulting hospitals were following:

- Routine USG check-up (18)
- Abdominal pain started at home (15)
- Does not felt foetal movement (3)
- Bleeding started at home (1)
- Bleeding with pain (2)
- Bleeding with water discharge (1)
- Scheduled LSCS due to IVF (1)
- Water discharge at home (2)
- Felt uneasy at home (1)
- Follow up suggested during previous visit (2)

Out of 46 total majority were visited health facility for routine USG examination and they come to know about Intra uterine foetal death and further management was proceed. Among 46 cases, 13 cases visited private facilities, 28 cases visited government facilities and remaining 5 cases visited trust hospitals.

Process in Private Health Facilities (N = 13)

Out of 13 cases visited private hospital, 12 cases were found IUFD on USG, while only 1 case had FHS present on admission and normal vaginal delivery induced but found stillborn as mother has bleeding prevaginal before labor to after delivery and mother was saved but baby didn’t survive. Remaining out of 12 cases, 8 cases were delivered through LSCS and 4 cases were delivered through Normal Vaginal Delivery. Out of 8 LSCS, one was twin delivery having female dead and male alive child were born.

#### Process in Trust Hospitals (N = 5)

Out of 5 cases visited trust hospital, 4 cases were found IUFD on USG, while only 1 case had FHS present on admission and normal vaginal delivery induced but foetus didn’t cry and FHS absent and declared stillborn. Remaining 4 cases, 3 cases were delivered through LSCS and 1 case was delivered through Normal Vaginal delivery.

#### Process in Government Health Facilities (N = 28)

Out of 28 cases visited Government hospital, 22 cases were found IUFD on USG, while only 6 cases had FHS present on admission, but among them 4 converted to IUFD during kept under observation in the hospital, while among remaining 2 cases, normal vaginal delivery induced. Baby was stuck during labor (“*Adadhu bahar ane adadhu andar – half in half outside “-25-year-old mother*) in one case and FHS were declining in second case and stillborn. Remaining 26 cases, 7 cases were delivered through LSCS and 19 cases was delivered through Normal Vaginal delivery.

#### Delivery conducted after ≥ 2 ^nd^ contacted hospitals (N = 12)

In this part, we discussed the care pathway of those cases who visited ≥ 2 Health facility just before delivery. We divided this part into 2 sub-groups based on decision taken for 2 ^nd^ or more facility visit: (a) Mother decided to shift to other hospital and (b) hospital referred to other hospital.

#### Hospital referred to other hospital (N = 9)

In this subgroup, cases with complaints visited the 1 ^st^ hospital – 6 cases in private hospitals and 3 cases in government hospital. But further they referred to other hospital by doctors to provide better service.

One case of abdominal pain, 3 cases of routine USG follow up and 2 cases of bleeding per vaginal visited private hospital initially and declared IUFD on USG. After that, one case referred to trust hospital and delivered through LSCS, while remaining 5 cases referred to government hospital, from which, two were delivered through LSCS. One delivery has twin birth having female child was still born and male child was alive. Remaining 3 cases referred to government hospital delivered through normal vaginal delivery.

One case of bleeding per vaginal, one case not feeling foetal movement and one case not feeling well consulted government hospital initially. Out of this three, one was alive, but due to bleeding was excessive referred to higher government centre, where lie management done to induce normal delivery. During this FHS drop rapidly and emergency LSCS performed but stillbirth happened.

while another one has inconclusive USG report and referred to higher centre to provide better service, but on approaching higher facility, they declared IUFD on USG and sent back to 1^st^ hospital, where normal delivery conducted. Last case was found IUFD on USG, but referred to higher centre where normal delivery conducted.

#### Mother decided to shift to other hospital (N = 3)

In this subgroup, cases with complaints visited the 1st hospital—2 cases in private hospitals and 1 case in a government hospital.

In the first case, the mother didn’t find a doctor during the initial hospital visit. They then went to a nearby 2nd private hospital, where IUFD was diagnosed on ultrasound investigation (USG). However, the mother refused to deliver there and returned to the 1st hospital, where they had received complete antenatal care. Despite waiting the entire night, the doctor didn’t arrive, so they shifted to another nearby private hospital, where a caesarean section (LSCS) was performed.

In the second case, a visit to a private hospital revealed IUFD on USG. The doctor recommended LSCS, but the mother declined permission. They then shifted to another nearby private hospital, where a normal delivery was conducted.

In the third case, the mother initially visited a nearby government hospital, where IUFD was diagnosed. However, due to fever, the mother was referred to a higher-level centre. Despite the referral, the family chose to visit a nearby private hospital, which declined the case. Finally, they complied with the referral and visited a higher-level government hospital, where a normal delivery took place.

### Objective 3: To identify associated factors and causes for stillbirth from community and health care provider perspectives

The following are the key finding based on qualitative analysis. There were two groups: (1) Health care provider and (2) Community

#### Health Care provider perspective

Total 17 Health care Providers - medical officer (2), ANM (13) and ASHA (2) were interviewed to understand associated factors and causes for stillbirths. Along with this, 9 family members were interviewed to understand community perspective related to factors and causes for stillbirth.

There were two basic themes: knowledge regarding classification of stillbirth and causes and factors related to stillbirths

##### Theme 1

knowledge regarding classification of stillbirth

##### Category 1

definition

We have taken stillbirth definition of “It is complete expulsion or extraction from a woman of a foetus following its death prior to the complete expulsion or extraction, at 28 or more completed weeks of gestation. When information on gestational age is unavailable use birthweight less than 1000 gm as the criteria” according to **ICD 10 classification of disease**. Based on this definition we divided knowledge of definition in two parts as per following: (Table 3)

**TABLE 3:**
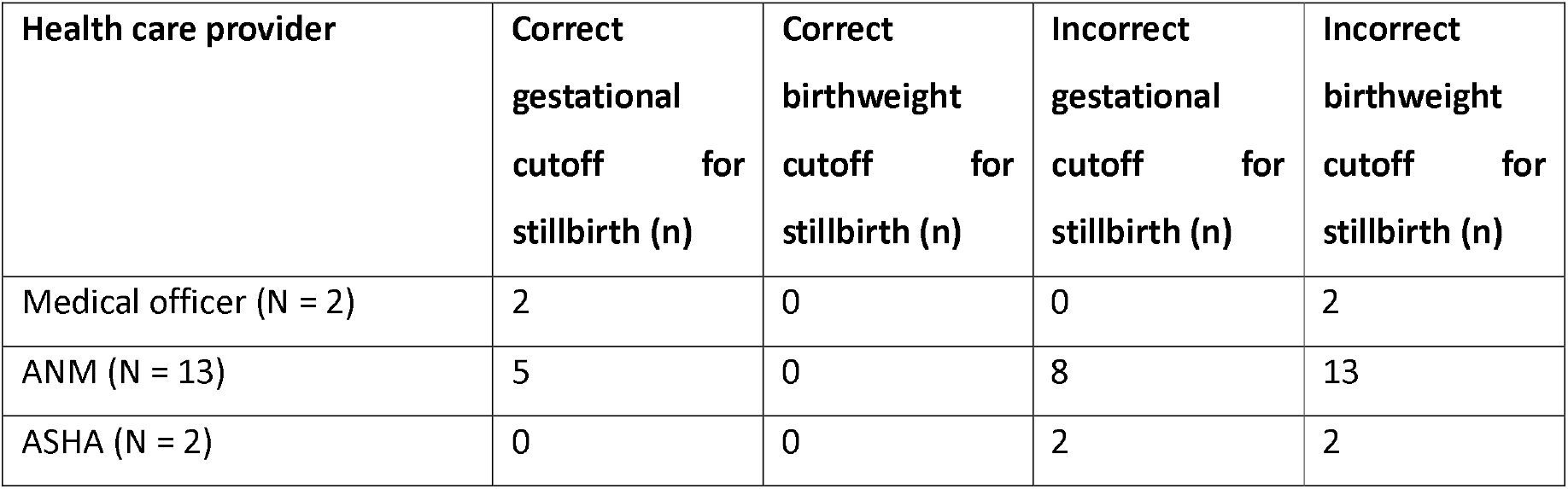
KNOWLEDGE OF HEALTH CARE PROVIDER RELATED DEFINITION OF STILLBIRTH (N = 17)

- 5 months of gestation and weight >900 gm (3 ANM),
- 7 months of gestation only (2 Medical officer and 5 ANM)
- 5 months of gestation only (5 ANM and 2 ASHA)

##### Category 2

training

We found that no training was provided to ANM or ASHA workers related to classification and verbal Autopsy filling related to stillbirths. Medical officers know the definition of stillbirth because they have studied it during MBBS.

##### Theme 2

causes and factors related to stillbirths

From the interview of health care provider, we enlisted following 12 causes and factors for stillbirth. Out of 12 reasons most common reasons were Premature delivery, Growth retardation, Hypertension, Diabetes, Unknown in top 5 of total list.(Table 4)

**TABLE 4:** FREE LISTING OF CAUSES AND FACTORS FOR STILLBIRTHS GIVEN BY HEALTH CARE PROVIDER (N = 17)

| Item | Frequency (%) | Average Rank | Salience |
| --- | --- | --- | --- |
| Premature delivery | 100 | 3.06 | 0.727 |
| Growth retardation | 88.2 | 3.53 | 0.59 |
| Hypertension | 82.4 | 2.86 | 0.637 |
| Diabetes | 82.4 | 3.93 | 0.523 |
| unknown | 82.4 | 5.86 | 0.293 |
| Meconium Aspiration | 76.5 | 3.92 | 0.466 |
| Low HB | 70.6 | 4.42 | 0.413 |
| twin pregnancy | 52.9 | 6.67 | 0.154 |
| Congenital defect | 47.1 | 7.13 | 0.117 |
| Birth Asphyxia | 35.3 | 4.67 | 0.192 |
| Placenta previa | 11.8 | 7 | 0.04 |
| Previous LSCS | 11.8 | 6 | 0.048 |

#### Pile sorting (Figure 3)

**FIGURE 3:**
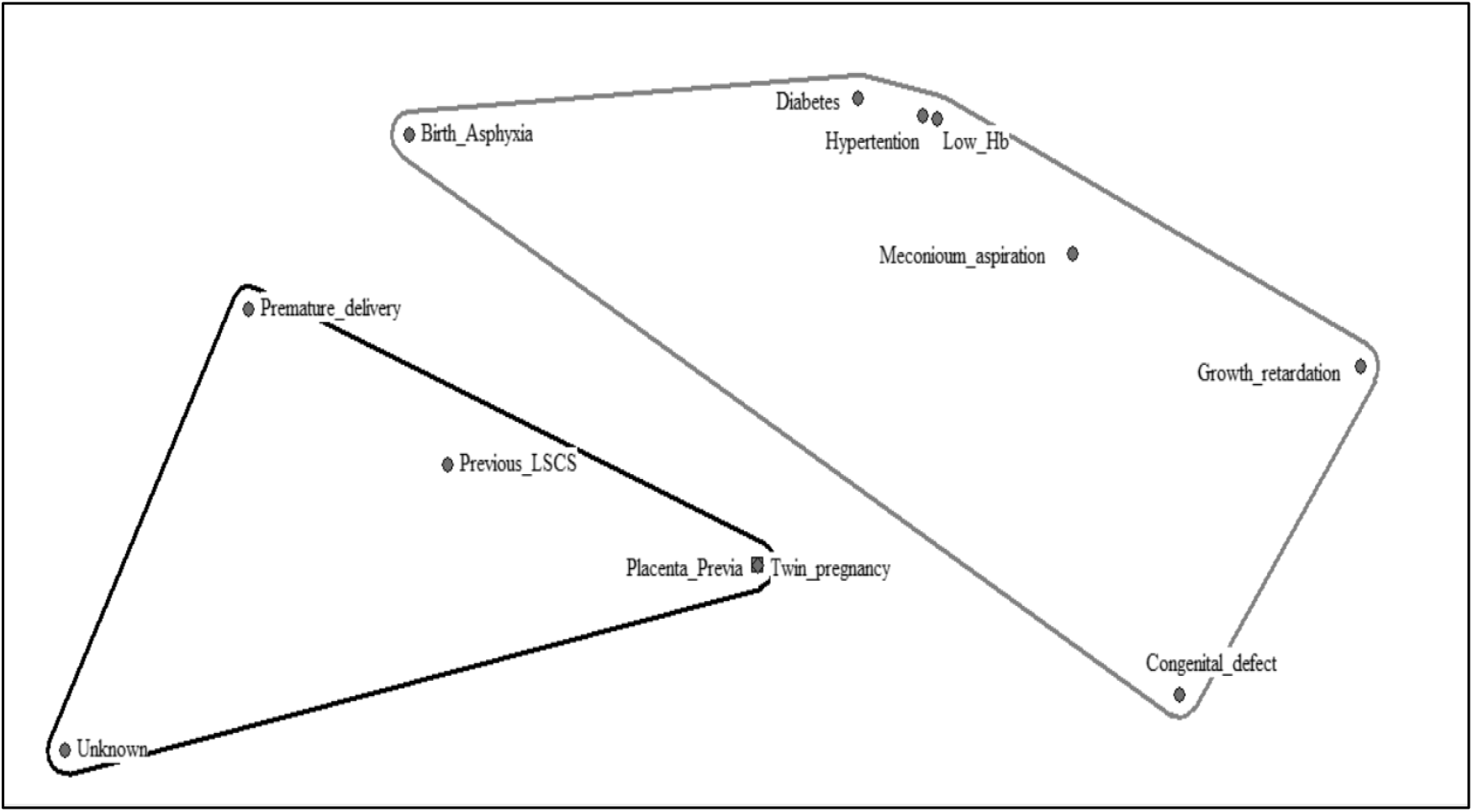
PILE SORTING OF CAUSES OF STILLBIRTH BASED ON PREVENTION (N = 17)

Based on pile sorting we identified two groups

##### Preventable stillbirth causes

Birth asphyxia, Diabetes and Hypertension during pregnancy, Low Hb, Meconium aspiration, Growth retardation and Congenital defect

##### Non-preventable causes

Premature Delivery, Previous LSCS, Placenta Previa, Twin Pregnancy, Unknown.

Based on these piles we can state that preventable causes highlight areas where targeted interventions and improved prenatal care can significantly reduce the incidence of stillbirths. For instance, **better management of maternal health conditions** like diabetes and hypertension, **ensuring adequate nutrition** to prevent low haemoglobin levels, and **monitoring foetal growth** can help in preventing these stillbirths. While non-preventable causes suggest that despite optimal care, some stillbirths may be unavoidable due to inherent medical conditions or complications. However, understanding these causes can help in preparing and managing high-risk pregnancies more effectively, potentially mitigating some risks.

#### Community perspective

Total 9 families who experienced stillbirth, were interviewed to understand associated factors and causes of stillbirth. The following are the findings based on qualitative analysis.

**There was one major theme:** factors and causes of stillbirths

Nine families were interviewed at their home after the stillbirth reporting communicated from VMC to investigator and following categories were emerge under major theme. The thematic analysis depicted in the Table 5 and Table 6 (supplementary file) illustrates various factors contributing to stillbirths, categorized into four main domains:

1. **Mother-related care**
2. **Facility-related care**
3. **Lack of financial support from government schemes**
4. **Medical conditions**. (Table 5)

**TABLE 5:**
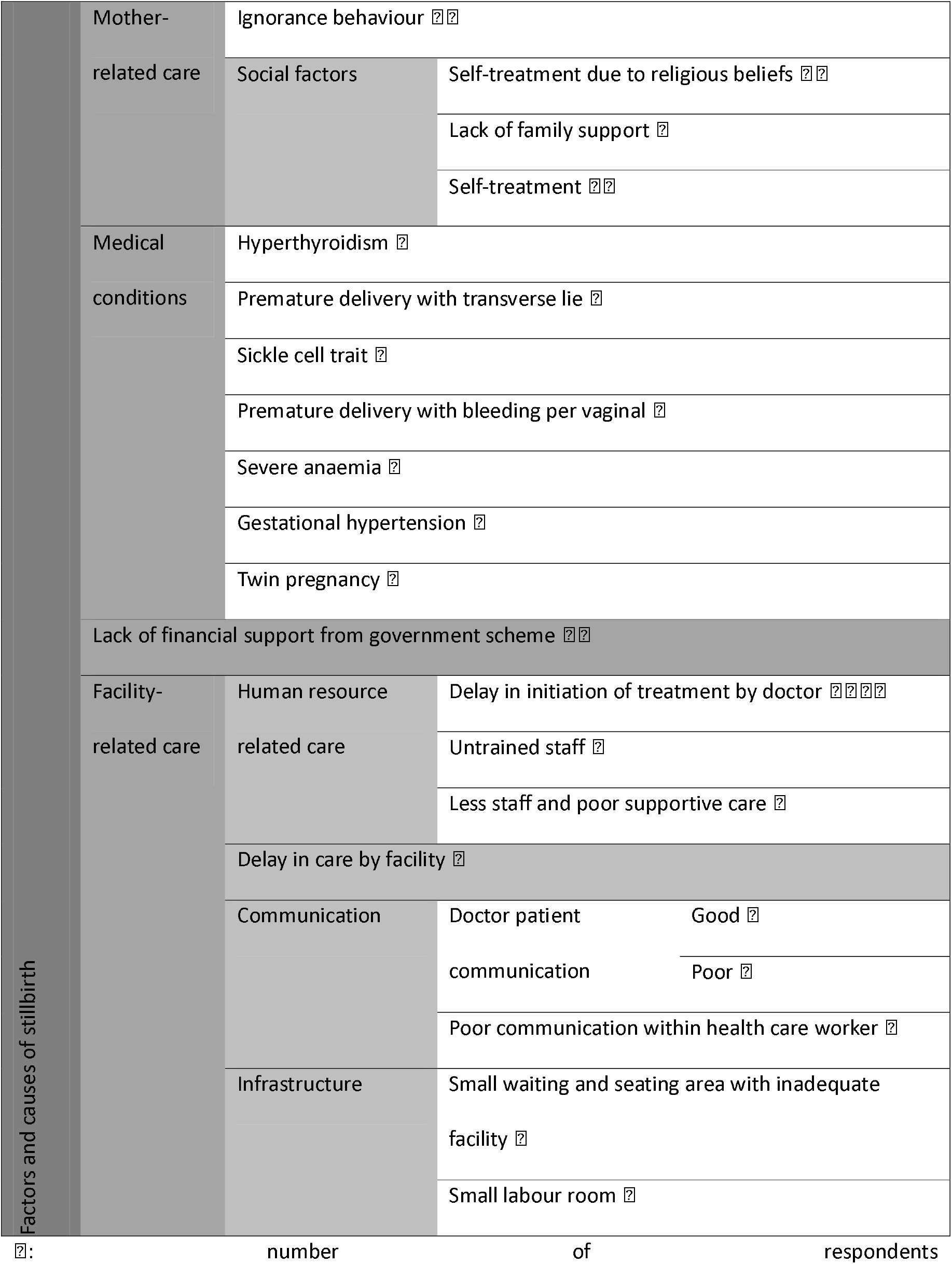
THEMATIC ANALYSIS OF FACTORS AND CAUSES OF STILLBIRTH BASED ON COMMUNITY PERSPECTIVE.

## DISCUSSION

After correcting for misreporting and data entry errors, the adjusted stillbirth rate (SBR) in urban Vadodara was 1.7 per 1000 total births. This rate is substantially lower than the national urban average of 3.0 and the Gujarat state average of 4.0 per 1000 total births reported by the Sample Registration System (SRS) 2020(Office of the Registrar General & Census Commissioner 2022), and successfully achieves the single-digit target set under the India Newborn Action Plan (INAP)(Ministry of Health & Family Welfare and Government of India 2014). However, the detection of seven reporting errors (11.5%)—including duplicate entries, misclassification of neonatal deaths and abortions as stillbirths, and gestational age miscalculations—reveals that the favourable rate is built upon a fragile data foundation, where nearly one in eight reported events is inaccurately documented.

The observed reporting inaccuracies correlate directly with systemic knowledge gaps identified among frontline providers. While medical officers correctly understood stillbirth definitions, most ANMs and ASHAs lacked clarity regarding the 28-week gestational or 1000g birthweight cutoffs, and none had received formal training on verbal autopsy classification. This disparity explains why errors in gestational age calculation (5.2% of cases) and gender misreporting occurred at the UPHC level, and suggests that the current HMIS figures may underestimate or misclassify the true burden, despite the low aggregate rate. The finding that 86.2% of stillbirths were antepartum, with intrauterine fatal death detected on routine ultrasound (25.9%) and premature delivery (19.0%) as the leading reported causes, aligns with the etiological patterns described by Newtonraj et al.(Newtonraj et al. 2017) and Tai et al.(Tai et al. 2021). Notably, the proportion of unexplained stillbirths in this study was only 6.9%, markedly lower than the 23–52.6% reported in other Indian settings(Kumbhare and Maitra 2016; Prasad et al. 2021). This discrepancy likely reflects the promptness of our verbal autopsy interviews (conducted within 21 days), which minimized recall bias and allowed families to articulate specific events, rather than a true biological difference in etiological distribution.

Geographically, the South zone recorded a rate of 4.1 per 1000—more than eight times higher than the West zone (0.5 per 1000)—indicating that the city-wide average masks profound intra-urban inequities. This variation is not explained by birth volume alone, as the East zone had the highest absolute number of stillbirths (n=21) but a lower rate (2.3 per 1000). Although the city relies heavily on government tertiary care (63.8% of all deliveries, with SSG Hospital alone accounting for 48.3% of stillbirth deliveries), the South zone paradoxically exhibited the highest rate (4.1 per 1000). This indicates that the presence of a major tertiary facility in the vicinity does not automatically translate to lower rates in that zone. The qualitative reports of delayed care initiation, undertrained staff, and overcrowded facilities—reported by families across all zones—provide a plausible systemic context for this disparity, although the study did not localize these themes exclusively to the South zone. Demographically, 10.3% of mothers belonged to high-risk age groups (<20 or >35 years), consistent with Prasad et al.(Prasad et al. 2021), who identified extreme maternal age as a significant predictor of intrauterine death. Additionally, 48.3% of affected women were primigravida and 53.4% were housewives, suggesting that first-time mothers with limited economic independence constitute a vulnerable subgroup where symptom recognition and timely care-seeking may be particularly compromised.

Clinically, conditions amenable to monitoring—such as hypertensive disorders (present in 19.0% of mothers during delivery), anaemia (12.1%), and meconium aspiration (6.9%)—were prominently cited by both families and healthcare providers. While Parmar et al.(Parmar et al. 2019) reported a higher anaemia burden in Saurashtra, our data show that even a 12.1% prevalence contributes to adverse outcomes in this setting, as evidenced by the 13.8% of stillbirths directly attributed to low birth weight and the qualitative theme of “pale and white appearance” observed in 5.2% of mothers. The identification of sickle cell trait in 3.4% of mothers, coupled with its association with stillbirth risk(Canelón et al. 2021), further underscores that locally prevalent genetic conditions are active contributors, even in an urban population with relatively high ANC coverage (93.1% underwent anomaly scans).

Care pathway analysis revealed that while 79.3% of deliveries were managed at the first contacted hospital, the remaining 20.7% involved multiple hospital visits—either through formal referrals or mother self-decision. In the referred subgroup, the initial diagnosis of IUFD at a private or trust hospital often led to onward referral to government facilities, where 55.6% ultimately underwent normal vaginal delivery. Critically, the qualitative narratives from families identified “delay in initiation of treatment by doctor” as the most frequently cited factor (reported by four families), alongside inadequate communication and “poor supportive care.” This fragmented pathway, where foetal heart sounds were present on admission in six government hospital cases but four later converted to IUFD under observation, points to a monitoring gap during the intrapartum period that is consistent with the provider-identified causes of birth asphyxia and meconium aspiration. Furthermore, the financial barriers related to accessing government scheme benefits (PMMVY – Pradhan Mantri Matru Vandana Yojana) reported by families highlight those administrative hurdles, rather than clinical complexity alone, may delay care-seeking, aligning with the 19.0% of cases presenting with premature rupture of membranes before labor onset.

## CONCLUSION

The review of stillbirth data from Vadodara Municipal Corporation identified discrepancies in reporting and classification, with 7 cases of misreporting and 2 data entry errors out of 61 entries. After corrections, the final list comprised 58 stillbirths, resulting in a stillbirth rate of 1.7 per 1000 total births, which is lower than the national and state averages. The study achieved the single-digit stillbirth rate goal set by INAP 2014, indicating a positive trend in stillbirth reduction within the corporation area.

The assessment of care pathways revealed that most families sought care at the first hospital they consulted, with a significant proportion of stillbirths detected during routine ultrasound investigations. A minority of families consulted multiple hospitals due to referrals or dissatisfaction with care, highlighting gaps in healthcare access and quality. Health problems experienced by families included maternal conditions such as anaemia, hypertension, and hyperthyroidism, emphasizing the need for comprehensive antenatal care to manage these factors and improve pregnancy outcomes.

Analysis from community and healthcare provider perspectives identified several associated factors and causes of stillbirth, including delays in care, untrained staff, inadequate infrastructure, and financial constraints. Mother-related factors such as ignorance of symptoms and reliance on self-treatment were also significant contributors. Medical conditions like severe anaemia, sickle cell trait, and gestational hypertension were common factors, underscoring the importance of targeted interventions, education, and improved healthcare services to reduce the incidence of stillbirths.

## Data Availability

All data produced in the present study are available upon reasonable request to the authors.

## RECOMMENDATIONS

- Improve reporting: Standardize training for ANMs/ASHAs on classification; conduct regular audits; digitize records.
- Enhance care pathways: Strengthen ANC access, referral systems, and infrastructure; address staffing and training gaps.
- Address risk factors: Target high-risk groups; improve management of anaemia, hypertension, and SCT.
- Community engagement: Increase awareness of warning signs, dangers of self-treatment, and importance of timely care.
- Financial access: Streamline government scheme processes.

## STRENGTHS OF THE STUDY

- Rare community-based cross-sectional study in urban area
- Complete population coverage minimizing sampling bias
- Multi-level healthcare institution inclusion
- Dual perspectives (families + providers)

## LIMITATIONS

- Urban setting limits rural generalizability
- Potential recall bias
- Retrospective classification challenges
- Non-standardized, non-validated verbal autopsy tool

## REFERENCES

Altijani N, Carson C, Choudhury SS, et al (2018) Stillbirth among women in nine states in India: rate and risk factors in study of 886,505 women from the annual health survey. BMJ Open 8:22583. 10.1136/BMJOPEN-2018-022583

Canelón SP, Butts S, Boland MR (2021) Evaluation of Stillbirth among Pregnant People with Sickle Cell Trait. JAMA Netw Open 4:. 10.1001/JAMANETWORKOPEN.2021.34274

Kumbhare SA, Maitra NK (2016) Aetiological Classification of Stillbirths: A Case Control Study. J Obstet Gynaecol India 66:420. 10.1007/S13224-015-0695-8

Lawn JE, Blencowe H, Waiswa P, et al (2016) Stillbirths: Rates, risk factors, and acceleration towards 2030. The Lancet 387:587–603. 10.1016/S0140-6736(15)00837-5

Ministry of Health & Family Welfare, Government of India (2014) INAP - India Newborn Action Plan

Newtonraj A, Kaur M, Gupta M, Kumar R (2017) Level, causes, and risk factors of stillbirth: A population-based case control study from Chandigarh, India. BMC Pregnancy Childbirth 17:1–9. 10.1186/S12884-017-1557-4/TABLES/3

Parmar MT, Karena Z V., Shah KD (2019) One year observational study of stillbirths in a referral hospital of Saurashtra region. Int J Reprod Contracept Obstet Gynecol 9:18. 10.18203/2320-1770.ijrcog20195572

Prasad AM, Agrawal M, Laddha A (2021) A CLINICAL STUDY OF RISK FACTORS AND MATERNAL OUTCOME IN INTRA-UTERINE FETAL DEATH. Int J Med Biomed Stud 5:. 10.32553/ijmbs.v5i8.2077

Singh S, Doyle P, Campbell OMR, Murthy GVS (2019) Management and referral for high-risk conditions and complications during the antenatal period: knowledge, practice and attitude survey of providers in rural public healthcare in two states of India. Reprod Health 16:. 10.1186/S12978-019-0765-Y

Tai NN, Mulchandani R, Modi DA, Rami B (2021) A prospective study of intrauterine death cases at a tertiary care hospital. Int J Reprod Contracept Obstet Gynecol 10:638. 10.18203/2320-1770.IJRCOG20210319

United Nations Inter-agency Group for Child Mortality Estimation (UN IGME) (2023) Never forgotten: the situation of stillbirth around the globe

World Health Organization (2019) QA46.1 Single stillbirth.In: International statistical classification of diseases and related health problems, 11th edn. World Health Organization

Office of the Registrar General & Census Commissioner I (ORGI) (2022) SAMPLE REGISTRATION SYSTEM (SRS)-STATISTICAL REPORT 2020. India

